# Evaluating COVID-19 screening strategies based on serological tests

**DOI:** 10.1101/2020.06.12.20129403

**Authors:** Michela Baccini, Alessandra Mattei, Emilia Rocco, Giulia Vannucci, Fabrizia Mealli

## Abstract

**Background:** Facing the SARS-CoV-2 epidemic requires intensive testing on the population to early identify and isolate infected subjects. Although RT-PCR is the most reliable technique to detect ongoing infections, serological tests are frequently proposed as tools in heterogeneous screening strategies. We analyze the performance of a screening strategy proposed in Tuscany (Italy), which first uses qualitative rapid tests for antibody detection, and then RT-PCR tests on the positive subjects.

**Methods:** We simulate the number of RT-PCR tests required by the screening strategy and the undetected ongoing infections in a pseudo-population of 500’000 subjects, under different prevalence scenarios and assuming a sensitivity of the serological test ranging from 0.50 to 0.80 (specificity=0.98). A compartmental model is used to predict the number of new infections generated by the false negatives two months after the screening, under different values of the infection reproduction number.

**Results:** Assuming a sensitivity equal to 0.80 and a prevalence of 0.3%, the screening procedure would require on average 11167.6 RT-PCR tests and would produce 300 false negatives, responsible after two months of a number of contagions ranging from 526 to 1132, under the optimistic scenario of a reproduction number between 0.5 to 1. Costs and false negatives increase with the prevalence.

**Conclusions:** The analyzed screening procedure should be avoided unless the prevalence and the rate of contagion are very low. The cost and effectiveness of the screening strategies should be evaluated in the actual context of the epidemic, accounting for the fact that it may change over time.

## INTRODUCTION

The severe acute respiratory syndrome coronavirus 2 (SARS-CoV-2) epidemic has rapidly spread around the world. Most European countries have implemented progressive measures of physical distancing. In Italy, from March 9^th^, citizens were prohibited from leaving home except in cases of proven need or urgency; since May 4^th^, the restrictions has been progressively lifted on. During this post lockdown period, scientists have advocated the need of implementing 3T-strategies, i.e., tracing, testing, treatment, in order to minimize the chance of a new spread of the disease and intercept as many infected individuals as possible [1]. The WHO has stressed that more intensive testing of suspected cases are required to identify and early quarantined infected people [2].

Knowledge of diagnostic tests for SARS-CoV-2 is still evolving, and a clear understanding of the nature of the tests and interpretation of their findings is not yet there. The most reliable diagnostic test for SARS-CoV-2 is the infections-reverse transcriptase-polymerase chain reaction (RT-PCR) test, although evidence arose that its accuracy could be not maximum [3-6].

A wide range of serology immunoassays (IAs) have been developed as well [7-8]. These include automated chemiluminescent IA (CLIA), manual ELISA, and rapid lateral flow IA (LFIA), which detect the immunoglobulin M (IgM) and immunoglobulin G (IgG) produced in persons in response to SARS-CoV-2 infection.

Due to the limited availability of reagents for RT-PCR tests and the relative low cost of the serological test, different countries and regions have proposed the use of IAs in combination with RT-PCR in heterogeneous screening strategies to detect subjects with ongoing infection, despite serological tests are not appropriate to reveal the presence of viral material during the infection. In fact, while they are useful to investigate the extent of the contagion in the community by detecting individuals who have developed antibodies, IAs may lead to high false negative and false positive rates if used with the intent of identifying subjects with ongoing infection, for reasons related to the SARS-CoV-2 antibody dynamics [6, 7, 9, 10].

In this paper, we analyze one such screening strategy, that has been proposed by the governor of Tuscany (Italy) with Decree n.54 of May 6^th^ 2020 [11]. This strategy, similar to others implemented elsewhere, uses first qualitative serological rapid tests, and then RT-PCR tests in case of positive immune response. The strategy is going to be applied to a large portion of the regional population: half a million people, approximately 1/8 of the whole population. Under different scenarios of prevalence of infection, we assess the performance of this screening strategy in terms of expected number of RT-PCR tests to be used on infected and uninfected subjects, as well as in terms of number of infected individuals that the procedure will not be able to detect. In order to contextualize the danger derived from the infected subjects left undetected, we quantify the potential contagion deriving from these false negatives under different hypotheses on the infection reproduction number, R_0_, that is, the average number of contagions deriving from one infected individual [12-13].

## METHODS

### Accuracy of the serological test if used to detect ongoing infection

Rapid point-of-care tests for detection of antibodies have been widely developed and marketed and are of variable quality. These tests are purely qualitative in nature and indicate the presence or absence of SARS-CoV-2 antibodies. A positive result may arise in case of 1) previous infection (high IgG titer); 2) infection in action (presence of both IgM and IgG); 3) none of the first two (false detection of antibodies). A negative test may arise in case of 1) early stage of an infection; 2) no previous infection; 3) false detection of absence of antibodies [7, 14].

Serological tests generally have a relatively high sensitivity as tests for detecting the presence of antibodies (IgG and IgM) [8]. However, due to reasons related to antibodies kinetics, infected individuals need some time to develop antibodies, and thus, serological tests wrongly report a negative result on infected individuals who have not developed antibodies, yet [6]. Therefore, serological tests are powerful diagnosis tools for asymptomatic patients or patients with mild to moderate illness who undergo the test after two weeks from the illness onset, but may perform poorly if used as screening tests for detecting ongoing infections [8].

The sensitivity of the serological test when used to detect ongoing infections can be decomposed in the following way:

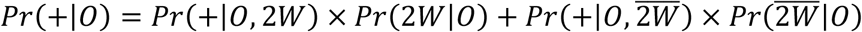

where, + = the serological test is positive, *O* = the subject has ongoing infection, *2W* = the test is performed during the first two weeks from infection onset. Let us assume that the probability that the serological test is positive in subjects that develop antibodies is at its maximum, 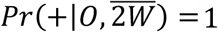, and that the time from infection onset to recovery is estimated as 4-6 weeks, so that *Pr*(2*W*|*O)* lies between 0.5 and 0.33[15]. Under this assumptions, if the probability of a positive serological test during the first two weeks of infection is equal to zero, *Pr*(+|*O*, 2*W*) = 0, then *Pr*(+|*O)* ranges from 0.5 to 0.67. If, more optimistically, *Pr*(+|*O*, 2*W)* is greater than zero, let say 0.30, then *Pr*(+|*O)* ranges from 0.65 to 0.77. It should be noticed that assuming 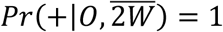 we get an optimistic range for the sensitivity *Pr*(+|*O*).

Regarding specificity, first note that each subject in the population belongs to one of the following disjoint groups: *O* = subjects who have ongoing infection; *Ī*= subjects who have never been infected; and *I*_*ō*_= subjects who have been infected in the past, but do not have an ongoing infection. Therefore, the event *Ō*, indicating that the subject does not have an ongoing infection, is equal to *Ī* ∪ *I*_*ō*_, and the event *I*, indicating that the subject has been infected, is equal to *O* ∪ *I*_*ō*_. Let – denote the event that the serological test is negative. Then, we can write the specificity of the serological test when used to detect ongoing infections, as follows:

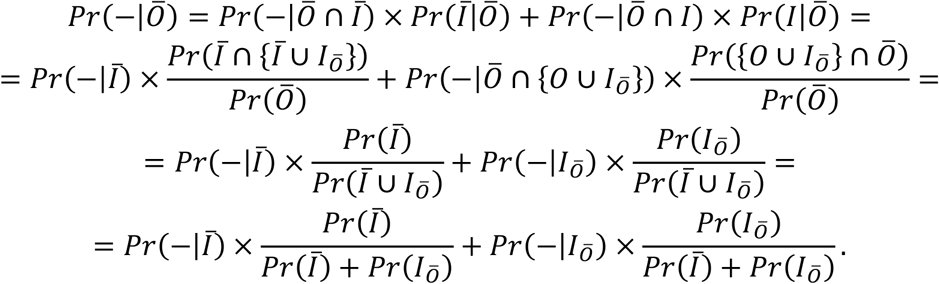

Under the assumption that the serological test is perfectly able to detect antibodies, unless it is performed within the 2 weeks window after the contagion, i.e. *Pr*(−|*Ō* ∩ *I*_*ō*_) = *Pr*(−|*I*_*ō*_) = 0, the previous equation becomes:

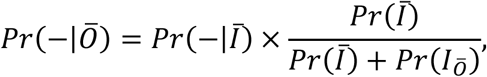

where *Pr*(−|*Ī*) is the specificity of the serological test if used to detect antibodies. Considering that *Pr*(−| *Ī*) is very high – approximately 1 for some serological tests [8, 16, 17] – and that in the actual epidemic scenario, being *Pr*(*Ī*) ≫ *Pr*(*I*_*O ō*_), the ratio *Pr*(*Ī*) *Pr*(*Ī*) + *Pr*(*I*_*O ō*_) is expected to be not lower than 0.98[12], in our analysis we set *Pr*(−| *Ō*) to 0.98.

### Monte Carlo simulation

We evaluate the performance of the screening strategy in a Monte Carlo simulation study on pseudo-populations of 500’000 subjects, characterized by different percentage of individuals with ongoing infection. We consider the following prevalence values: 0.003, 0.005, 0.010, 0.015, 0.02. The first three scenarios are in accordance with studies reporting proportions of infected peoples around 0.3% and never greater than 1% over the last four months of the coronavirus pandemic[12, 18], the last two correspond to scenarios where the level of contagion is worse than the one expected in the general population.

According to the results obtained in the previous section, we first focus on an optimistic scenario where sensitivity= 0.80 and specificity=0.98. Then, taking 0.80 as an upper bound of the sensitivity level, we also perform simulations with sensitivity equal to 0.5, 0.6 and 0.7.

For sake of simplicity, we assume that the RT-PCR test has specificity and sensitivity equal to 1, even if a certain percentage of false negatives is expected from this procedure as well [4-5]. Under this assumption, the sensitivity and specificity of the serological test can be interpreted as “relative” to the RT-PCR test.

For each simulation setting, we run 500 Monte Carlo (MC) iterations and in each iteration we calculate various performance measures of the strategy: total number of RT-PCR tests, number of RT-PCR tests respectively performed on infected and uninfected individuals, number of false negatives, and negative predictive value. It is worth noting that, due to the fact that (false and true) positives from the serological test undergo a RT-PCR test, the screening protocol does not produce false positives (the specificity of the strategy is 1 by design), thus the positive predictive value is 1. Starting from the estimated numbers of false negatives arising from the Monte Carlo simulations, we use a Susceptible-Infected-Recovered (SIR) compartmental model to predict the number of infected originated by those that the screening left undetected. Assuming a time from infection to exit (death or recovery) equal to three weeks [15, 19], we calculate both the number of circulating subjects with ongoing infection (assuming that all subjects in the population are screened at the same time) and the cumulative number of new infections two months after the screening, derived from the undetected infected subjects. Calculations are done under the different prevalence scenarios used in the MC simulations and different hypothetical values of the infection reproduction number, R_0_: 0.5, 1, 1.5, 2.

## RESULTS

In relative terms, the strategy appears to perform well: the negative predictive values are very high, with a MC mean always close to 1 and a very low variability as described by the 5% and 95% MC percentiles (see Figure 4). However, the actual impact can be better understood by looking at the absolute values: the total number of RT-PCR tests performed at the second step of the screening procedure and the number of false negatives.

Figure 1 shows the MC average number of RT-PCR tests (total number of RT-PCR tests on infected and uninfected individuals) and the average number of false negatives. In addition to the MC mean, Table 1 shows the MC 5^th^ and 95^th^ percentile of these quantities.

**Table 1.** Monte Carlo mean and 5th and 95th percentiles of the total number of RT-PCR tests, the number of RT-PCR tests on uninfected individuals, and the number of false negatives by prevalence of infection in the population and sensitivity of the serological test (specificity of the serological test=0.98).

| Prevalence | Number of RT-PCR tests |  |  | Number of RT-PCR on uninfected individuals |  |  | Number of False Negatives |  |  |
| --- | --- | --- | --- | --- | --- | --- | --- | --- | --- |
|  | Mean | 5 % | 95 % | Mean | 5 % | 95 % | Mean | 5 % | 95 % |
| *Specificity=0.98 and Sensitivity=0.50* |  |  |  |  |  |  |  |  |  |
| 0.003 | 10719.3 | 10557.0 | 10893.0 | 9967.6 | 9797.0 | 10139.0 | 748.3 | 715.0 | 778.0 |
| 0.005 | 11200.5 | 11029.9 | 11379.1 | 9947.5 | 9779.9 | 10117.0 | 1247.1 | 1205.0 | 1288.0 |
| 0.010 | 12401.6 | 12230.0 | 12588.1 | 9897.1 | 9736.4 | 10069.0 | 2495.5 | 2435.0 | 2549.0 |
| 0.015 | 13600.6 | 13420.0 | 13781.1 | 9847.0 | 9681.8 | 10018.0 | 3746.5 | 3671.0 | 3816.0 |
| 0.020 | 14801.8 | 14606.0 | 14988.2 | 9796.7 | 9628.0 | 9964.2 | 4994.9 | 4911.0 | 5075.0 |
| *Specificity=0.98 and Sensitivity=0.60* |  |  |  |  |  |  |  |  |  |
| 0.003 | 10866.8 | 10692.7 | 11038.1 | 9967.6 | 9797.0 | 10139.0 | 600.8 | 572.0 | 631.0 |
| 0.005 | 11446.6 | 11272.0 | 11614.1 | 9947.5 | 9779.9 | 10117.0 | 1000.9 | 961.9 | 1042.0 |
| 0.010 | 12894.5 | 12727.0 | 13066.1 | 9897.1 | 9736.4 | 10069.0 | 2002.6 | 1949.9 | 2061.1 |
| 0.015 | 14344.7 | 14158.0 | 14521.0 | 9847.0 | 9681.8 | 10018.0 | 3002.4 | 2934.0 | 3072.1 |
| 0.020 | 15793.1 | 15614.0 | 15964.4 | 9796.7 | 9628.0 | 9964.2 | 4003.7 | 3921.0 | 4085.0 |
| *Specificity=0.98 and Sensitivity=0.70* |  |  |  |  |  |  |  |  |  |
| 0.003 | 11017.3 | 10844.0 | 11188.0 | 9967.6 | 9797.0 | 10139.0 | 450.3 | 420.0 | 480.0 |
| 0.005 | 11697.1 | 11526.9 | 11865.1 | 9947.5 | 9779.9 | 10117.0 | 750.4 | 711.0 | 793.0 |
| 0.010 | 13396.1 | 13222.8 | 13570.2 | 9897.1 | 9736.4 | 10069.0 | 1501.0 | 1445.9 | 1556.0 |
| 0.015 | 15096.7 | 14906.9 | 15271.0 | 9847.0 | 9681.8 | 10018.0 | 2250.4 | 2180.9 | 2315.1 |
| 0.020 | 16795.7 | 16611.0 | 16976.0 | 9796.7 | 9628.0 | 9964.2 | 3001.1 | 2925.0 | 3079.0 |
| *Specificity=0.98 and Sensitivity=0.80* |  |  |  |  |  |  |  |  |  |
| 0.003 | 11167.6 | 10991.9 | 11333.1 | 9967.6 | 9797.0 | 10139.0 | 300.0 | 273.9 | 326.0 |
| 0.005 | 11947.2 | 11774.0 | 12111.1 | 9947.5 | 9779.9 | 10117.0 | 500.3 | 465.9 | 535.0 |
| 0.010 | 13896.8 | 13719.9 | 14067.2 | 9897.1 | 9736.4 | 10069.0 | 1000.4 | 952.0 | 1047.0 |
| 0.015 | 15846.6 | 15674.8 | 16030.1 | 9847.0 | 9681.8 | 10018.0 | 1500.5 | 1447.0 | 1556.0 |
| 0.020 | 17795.9 | 17625.7 | 17979.2 | 9796.7 | 9628.0 | 9964.2 | 2000.8 | 1935.0 | 2072.0 |

**Figure 1.**
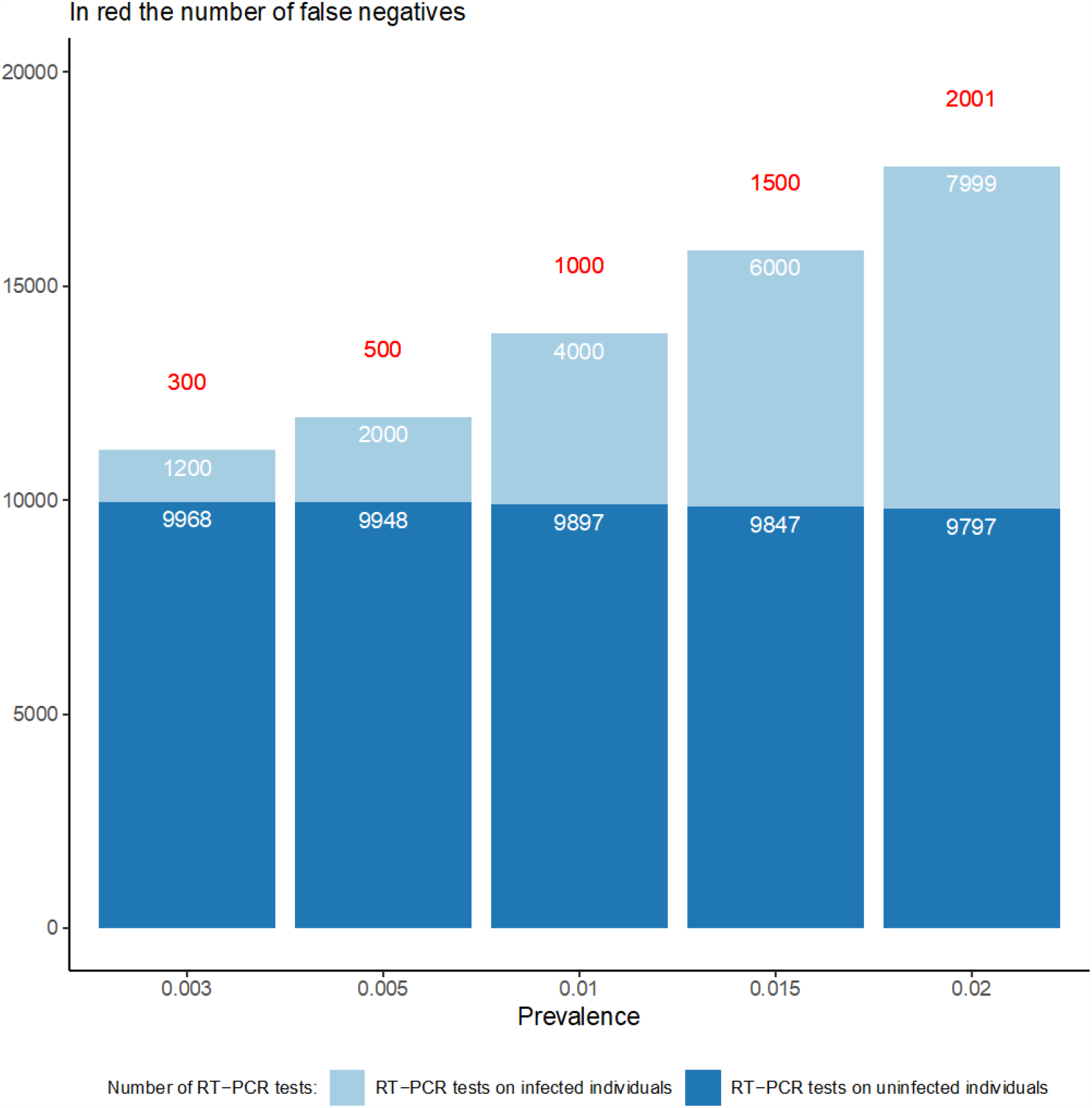
Number of RT-PCR tests on infected and uninfected subjects and number of false negative arising from the screening procedure, by prevalence of infection in the population (sensitivity of the serological test=0.80, specificity of the serological test=0.98).

For instance, consider a population where the prevalence is 0.3% and the specificity and sensitivity of the serological test are, respectively, 0.98 and 0.80 (first bar in Figure 1 and first row in Table 1). Two important findings are noted: First, a large number of RT-PCR tests is performed on uninfected individuals: the screening strategy requires that, in addition to the 500’000 serological tests, about 11’168 RT-PCR tests are administered, 9’968 of which undergone on uninfected people, that is, on false positive people to the serological test. Only the remaining 1’200 tests reveals the presence of the infection. Second, the screening strategy leads to 300 false negatives, i.e., the serological test is not able to detect the presence of antibodies in 300 infected individuals, possibly because done within two weeks from the onset of the infection. No test will be performed on these infected individuals, who, therefore, will not be kept in quarantine.

As the prevalence increases (Figure 1 and Table 1) the number of false negatives increases, too. On the other hand, the number of RT-PCR tests performed on the false positives slightly decreases. Leaving the specificity of the serological test unchanged, Figure 2 reports the expected number of false negatives as the sensitivity varies, for different values of prevalence.

**Figure 2.**
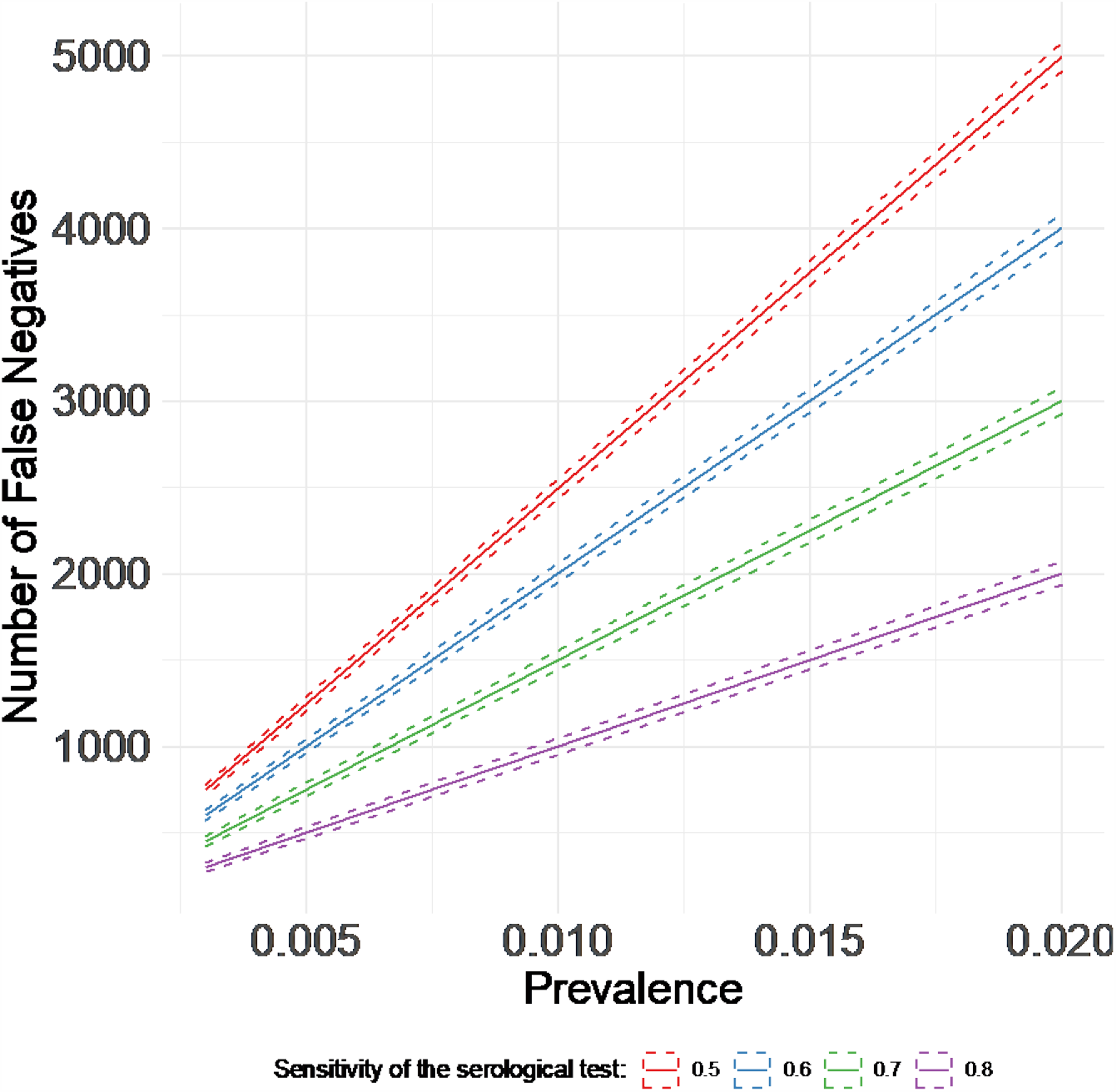
Number of false negatives arising from the screening procedure, by sensitivity of the serological test and prevalence of infection in the population (specificity of the serological test=0.98).

The results of the SIRD model are reported in Table 2 and Figure 3. For this analysis we assume a sensitivity of the test equal to 0.80 and a specificity equal to 0.98. In Figure 3 we report the daily number of infected derived from the false negatives, who is expected to be present in the region during the first month after the screening. The number of infected subjects decreases over time if the reproduction number, R_0_, is lower than 1, is stable if R_0_ is exactly 1 and increases if R_0_ is higher than 1. The induced epidemic strongly depends on the initial number of false negatives, and thus on the prevalence of infection at the time of the screening.

**Table 2.**
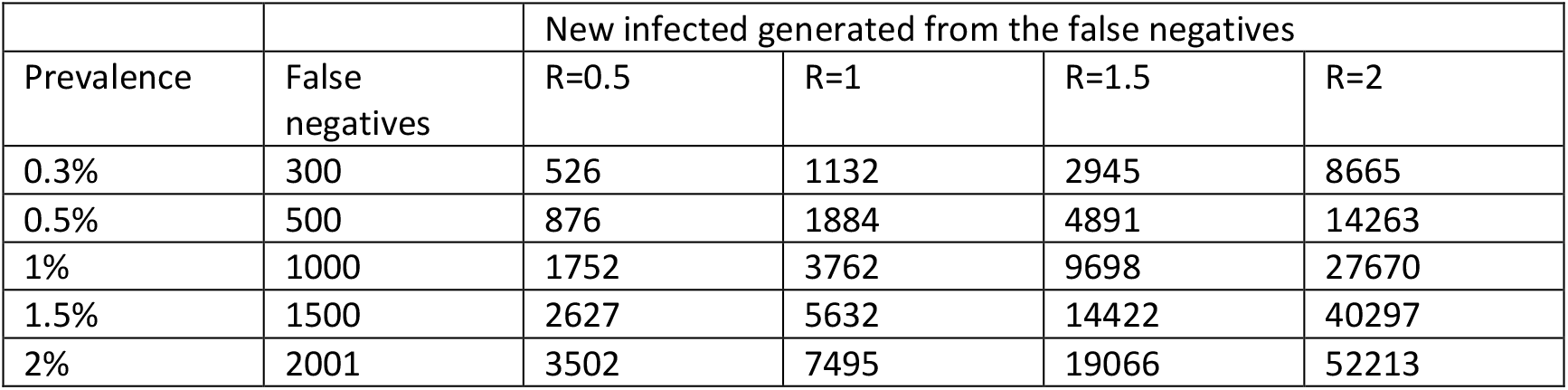
Cumulative number of new infected induced by the false negatives after two months from the screening, by prevalence of infection in the population and infection reproduction number (sensitivity of the serological test=0.80, specificity of the serological test=0.98).

**Figure 3.**
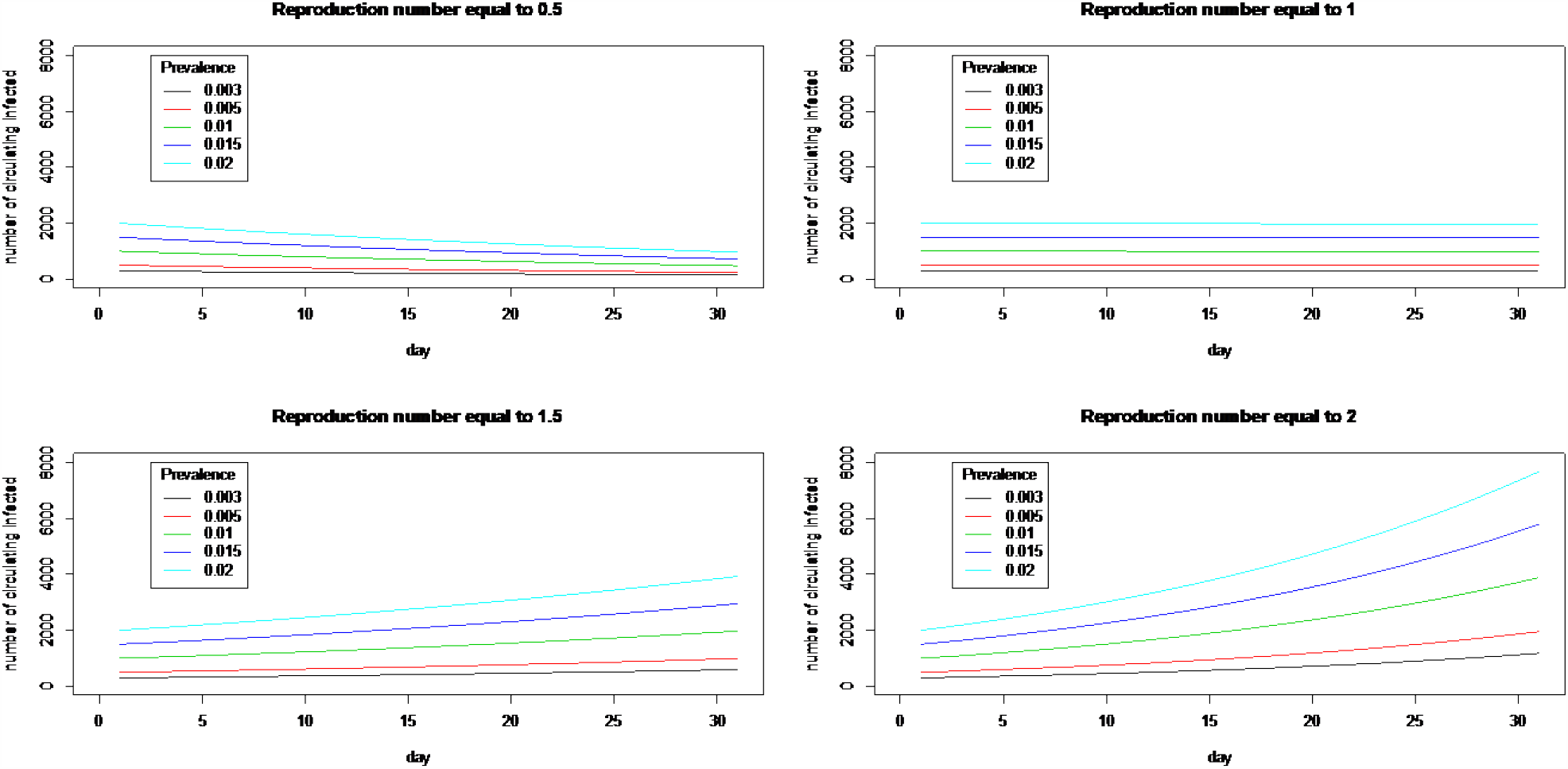
Number of infected subjects induced by the false negative to the serological test, circulating in the region during the first 30 days after the screening, by prevalence of infection in the population and infection reproduction number.

**Figure 4.**
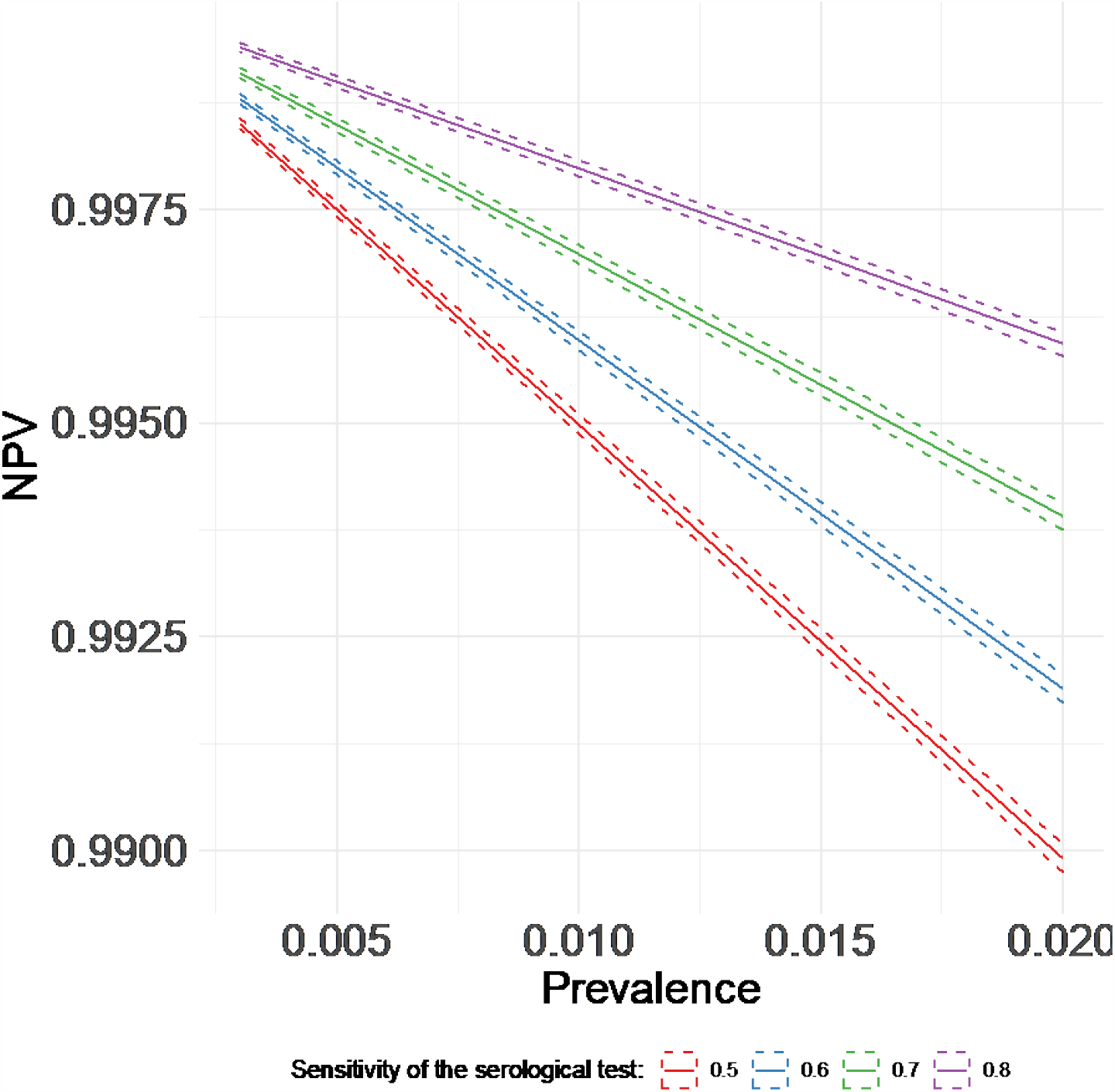
Negative predictive values arising from the screening procedure, by sensitivity of the serological test and prevalence of infection in the population (specificity of the serological test=0.98).

The total number of new infections attributable to the false negatives after two months from the screening is reported in Table 2. If R_0_ is lower than 0.5 and the prevalence is lower than 1%, the new infected attributable to the original false negatives are less than 1’800. But a values of R_0_ equal to 1 is sufficient to double the burden of infection originated from the undetected infected individuals. If the reproduction number is equal to 2, the new infections are expected to be more than 8’000, even for very low values of the initial prevalence.

## DISCUSSION

Our results provide important insights on the cost-effectiveness of the analyzed strategy. First of all, in a time of scarce availability of reagents for RT-PCR analysis, it should be carefully evaluated whether it is feasible to perform such a large number of RT-PCR tests on subjects resulted positive to the rapid serological test, by also considering that most of these positives are false positives. The fact that, as described in the Decree [11], serological tests will be carried out on a voluntary basis further undermines the performance of the strategy. Alternative less laborious and less costly screening procedures and strategies that improve the accuracy of the serological tests should be evaluated. For instance, in order to increase the positive predictive value of the serological test, a questionnaire collecting information on SARS-CoV-2 symptoms could be firstly administered and individuals with SARS-CoV-2 symptoms encouraged to undergo the test. Additionally, with the aim to reduce the costs related to the RT-PCR tests performed at the second step of the screening procedure, individuals with a positive serological result might be preventively quarantined and undergone a second serological test after a pre-fixed time period, defined on the basis of SARS-CoV-2 antibodies kinetics. In this case, social and economic costs deriving from the preventive quarantine should be quantified as well.

From an efficacy perspective, our results show that the main pitfall of the strategy is the number of undetected infected people, which increases as the prevalence of subjects with ongoing infection in the population increases. False negatives could cause false reassurance, behavioral changes, and disease spread. It is also important that the number of false negatives is evaluated in the context of the epidemic. In fact, each undetected case can generate other infected individuals and the transmission can happen with different strength, depending on many factors mainly related to the implementation of measures of physical distancing and plans aimed to early detect and isolate new cases. In our simplified SIR model [13], we change R_0_ to define different scenarios of transmission, showing that the same number of initial undetected infected individuals can be responsible of very different numbers of new infected individuals after two months from the screening. In particular, strategies like the one analyzed in this paper might be particularly ineffectiveness in the long-run, when, with the progressive time from the lockdown, an increase of R_0_ is expected, or when applied to sub-populations where the number of potential at risk contacts is higher than among the general population, such as patients or workers of hospitals and nursing homes. In addition, it should be also considered that the relative gain of applying this two-step screening procedure in respect to testing all subjects through individual RT-PCR tests decreases with the prevalence, and that, as the prevalence increases, the number of false negatives increases as well. Thus, this kind of procedure should absolutely be avoided unless the prevalence is very low.

In the context of the SARS-CoV-2 pandemic, evaluation procedures such as that one used in this paper should be routinely performed before any implementation of new screening strategies on the general population or on specific subpopulations, particularly in the presence of resources constraints. Unlike in the case of non-communicable diseases, the danger deriving from cases that are not detected by the screening should be contextualized accounting for the strength of the epidemic spread, which depends not only on the social distancing measures adopted to slow the contagion, but also on the measures undertaken for early detection and isolation of the subjects with ongoing infection, thus ultimately on the effectiveness of the screening strategies themselves.

## Data Availability

The manuscript results are based on simulations.

## Acknowledgments

We would like to thanks the working group on COVID-19 at the Department of Statistics, Computer Science, Application “G. Parenti” of the University of Florence, and specifically Carla Rampichini, Anna Gottard, Matteo Pedone, Fiammetta Menchetti, Silvia Noirjean, Cecilia Viscardi, Giulia Cereda.

## References

[1] Vespignani A. (2020) Per sconfiggere il virus servono le 3T. https://www.reccom.org/2020/05/04/vespignani-per-sconfiggere-il-virus-servono-le-3t/ (accessed 4 May 2020).

[2] WHO. (2020) Strategies and planning. https://www.who.int/emergencies/diseases/novel-coronavirus-2019/strategies-and-plans

[3] Lippi G, Simundic AM, Plebani M. Potential preanalytical and analytical vulnerabilities in the laboratory diagnosis of coronavirus disease 2019 (COVID-19) [published online ahead of print, 2020 Mar 16]. Clin Chem Lab Med. 2020;/j/cclm.ahead-of-print/cclm-2020-0285/cclm-2020-0285.xml. doi:10.1515/cclm-2020-0285.

[4] Abdalhamid B, Bilder CR, McCutchen EL, et al. Assessment of Specimen Pooling to Conserve SARS CoV-2 Testing Resources. Am J Clin Pathol. 2020;153(6):715–718. doi:10.1093/ajcp/aqaa064.

[5] He JL, Luo L, Luo ZD, et al. Diagnostic performance between CT and initial real-time RT-PCR for clinically suspected 2019 coronavirus disease (COVID-19) patients outside Wuhan, China. Respir Med. 2020;168:105980. doi:10.1016/j.rmed.2020.105980.

[6] Sethuraman N, Jeremiah SS, Ryo A. Interpreting Diagnostic Tests for SARS-CoV-2 [published online ahead of print, 2020 May 6]. JAMA. 2020;10.1001/jama.2020.8259. doi:10.1001/jama.2020.8259

[7] Italian Ministry of Health. Test di screening e diagnostici. http://www.normativasanitaria.it/jsp/dettaglio.jsp?id=74021 (accessed 9 May 2020).

[8] The Johns Hopkins Center for Health Security. Serology-based tests for COVID-19. https://www.centerforhealthsecurity.org/resources/COVID-19/serology/Serology-based-tests-for-COVID-19.html

[9] Watson J, Whiting PF, Brush JE. Interpreting a covid-19 test result. BMJ. 2020;369:m1808. Published 2020 May 12. doi:10.1136/bmj.m1808.

[10] Haugh T, Suneal B. Just because you test positive for antibodies doesn’t mean you have them. https://www.nytimes.com/2020/05/13/opinion/antibody-test-accuracy.html (5 May 2020).

[11] Decree n. 54 of May 6th 2020. https://www.cicogninirodariprato.edu.it/wp-content/uploads/2020/05/ordinanza_del_presidente_n-54_del_06-05-2020.pdf

[12] Baccini M, Cereda G, Viscardi C, et al. Oltre il picco: analisi e previsioni dell’epidemia da sars-cov-2 in Toscana. Epidemiologia &Prevenzione. https://repo.epiprev.it/index.php/2020/04/10/oltre-il-picco-analisi-e-previsioni-dellepidemia-da-sars-cov-2-in-toscana/ (10 mApr 2020) Repository: repo.epiprev.it/1085

[13] Lin F, Muthuraman K, Lawley M. An optimal control theory approach to non-pharmaceutical interventions. BMC Infect Dis. 2010;10:32. Published 2010 Feb 19. doi:10.1186/1471-2334-10-32.

[14] Li Z, Yi Y, Luo X, et al. Development and clinical application of a rapid IgM-IgG combined antibody test for SARS-CoV-2 infection diagnosis [published online ahead of print, 2020 Feb 27]. J Med Virol. 2020;10.1002/jmv.25727. doi:10.1002/jmv.25727.

[15] Who – China. https://www.who.int/china

[16] Corriere della sera – Brescia. https://brescia.corriere.it/notizie/cronaca/20_maggio_23/test-sierologici-622percentodegli-isolati-ha-anticorpiuno-5-ancora-infetto-cc3e316a-9cda-11ea-a31e-977f755d9d62.shtml (accessed 23 May 2020).

[17] Corriere della sera. https://www.corriere.it/salute/malattie_infettive/20_maggio_02/coronavirus-quanto-sono-davvero-accurati-test-sierologici-01ec7db0-8c52-11ea-9e0f-452c0463a855.shtml (accessed May 2020).

[18] Woodhill N. Coronavirus (covid-19) infection survey pilot: England, 4 May 2020. Statistical bulletin of Office for National Statistics. https://www.ons.gov.uk/peoplepopulationandcommunity/healthandsocialcare/conditionsanddiseases/bulletins/coronaviruscovid19infectionsurveypilot/england14may2020

[19] Istituto Superiore di Sanità. https://www.epicentro.iss.it/coronavirus/

